# Flu-CNN: predicting host tropism of influenza A viruses via character-level convolutional networks

**DOI:** 10.1101/2023.08.28.23294703

**Authors:** Nan Luo, Xin Wang, Boqian Wang, Renjie Meng, Yunxiang Zhao, Zili Chai, Yuan Jin, Junjie Yue, Mingda Hu, Wei Chen, Hongguang Ren

## Abstract

Throughout history, Influenza A viruses (IAVs) have caused significant harm and catastrophic pandemics. The presence of host barriers results in viral host tropism, where infected hosts are subject to strict restrictions due to the hindered spread of viruses across hosts. Therefore, the identification of host tropism of IAVs, particularly in humans, is crucial to preventing the cross-host transmission of avian viruses and their outbreaks in humans. Nevertheless, efficiently and effectively identifying host tropism, especially for early host susceptibility warnings based on viral genome sequences during outbreak onset, remains challenging. To address this challenge, we propose Flu-CNN, a deep neural network model based on classical character-level convolutional networks. By analyzing the genomic segments of IAVs, Flu-CNN can accurately identify the host tropism, with a particular focus on avian influenza viruses that may infect humans. According to our experimental evaluations, Flu-CNN achieved an accuracy of 99% in identifying virus hosts via only a single genomic segment, even for subtypes with a relatively small number of viral strains such as H5N1, H7N9, and H9N2. The superiority of Flu-CNN demonstrates its effectiveness in screening for critical amino acid mutations, which is important to host adaptation, and zoonotic risk prediction of viral strains. Flu-CNN is a valuable tool for identifying evolutionary characterization, monitoring potential outbreaks, and preventing epidemical spreads of IAVs, which contribute to the effective surveillance of influenza A viruses.

## 1 Introduction

Influenza A virus (IAV) is capable of infecting a wide range of hosts, including humans, birds, and other mammals [1]. Throughout history, IAV has become a frequent and leading cause of respiratory infections in both human and avian species, which may result in significant morbidity and mortality [2]. For human beings, IAV has caused several pandemics throughout history, among which the 1918-19 H1N1 influenza pandemic stands out, as it resulted in the deaths of nearly 50 million people and inflicted significant damage upon human health and well-being [3]. In terms of birds, the H5N1 avian influenza viruses have swept through Asia, Africa, Europe and North America since 2021, leading to the death of millions of poultry and wild birds [4]. To date, IAVs have caused several pandemics and have become a major and persistent threat to human and avian health.

One phenomenon is that IAVs can only infect specific hosts, which indicates that the IAV is restricted by its host tropism, i.e., host specificity [5]. This implies that IAVs have the adaptability of hosts [6]. The host tropism of IAVs is due to the presence of host barriers, which typically impedes the easy spread of these viruses across hosts. Consequently, avian influenza viruses are prevented from causing disease in humans by host barriers. However, IAVs may break host barriers through evolution, by acquiring mutations and reassortments that alter their receptor binding affinity and antigenicity [7, 8]. Some avian influenza viruses, such as H5N1, H7N9, and H9N2, have been reported to infect humans occasionally [9-11]. From January 2003 to April 2023, 868 cases of human infection with H5N1 avian influenza have been reported from 21 countries, including 457 deaths [12]. Therefore, these subtypes can be greatly dangerous to human beings. Meanwhile, phylogenetic studies have shown that the genes in waterfowl are often considered to be the origin of IAVs from other species [13]. This suggests that the changes in host tropism may be the main cause of cross-species transmission. Although the host barrier can protect humans from avian influenza viruses to some extent, avian influenza viruses can still pose a great threat to human health once they change their host tropism. Therefore, predicting host tropism of IAVs is of great importance in the surveillance of pandemics, especially for monitoring the cross-species transmission of IAVs.

Previous experimental studies have identified numerous factors that influence the host tropism of IAVs, including receptor binding affinity, viral genome replication, and host immunity antagonization [14-16]. However, it is still difficult to determine the host tropism of large numbers of IAVs efficiently and effectively, using experimental methods. Besides, performing such biological experiments also requires high standards of biological laboratories, which limits the scalability of such investigations. Therefore, many computational tools have been developed to analyze viral sequences for their host tropism, including distinct host tropism protein signatures, zoonotic risk of IAVs, avian influenza transmission from avian to human, and prediction of human-adapted IAVs based on viral nucleotide composition [17-19]. Most of these methods can be effective, but they require feature extraction from the input sequence and even particular analysis of host genomic information, which may limit their application. Furthermore, current methods may not fully leverage the vast amount of genomic data available for IAVs, resulting in the potential loss of critical information during the analysis process. Therefore, the performance and application of those methods may be greatly limited.

In this study, we propose a novel approach using Character-level Convolutional Neural Networks (Char-CNN) [20]. Inspired by classical Char-CNN models, our method analyzes the whole genome or some segments of IAVs to predict the viral host tropism. We had collected a large-scale dataset from three major databases, including NCBI Virus (https://www.ncbi.nlm.nih.gov/labs/virus) [21], GISAID (https://www.gisaid.org) [22], and BV-BRC (https://www.bv-brc.org) [23], which comprises both human and avian categories for model training and evaluation. To our knowledge, this is the first work which has used such a large-scale dataset for IAVs host tropism prediction. The evaluation result demonstrates that our approach can effectively identify the host tropism of IAVs with an accuracy rate of 99%, using just a single genomic segment of IAV. Moreover, our method can likewise achieve a stable and high accuracy across various subtypes, particularly on avian influenza subtypes with a small number of viral strains such as H5N1, H7N9, and H9N2. Furthermore, we have also explored our method in other perspectives. We have investigated the interpretability of Flu-CNN, and our method can learn the key features to distinguish the hosts by convolution. We also use Flu-CNN to explore the important amino acid substitutions which can change the IAV adaptation. Based on Flu-CNN, we have screened on PB2, PA (polymerase acidic protein) and NP (nucleoprotein) proteins to obtain some key amino acid substitutions. Moreover, we use Flu-CNN to identify the zoonotic risk of IAVs strains for estimating the potential high-risk strains circulating in avian. Our result demonstrates that H5N1, H7N9, and H9N2 subtypes have the highest zoonotic risk. This research produces a valuable tool for identifying the host tropism of IAV as well as innovative insights into the evolutionary characterization of IAV, which may contribute to the surveillance of potential outbreaks and spread of IAVs.

## 2 Materials and Methods

### 2.1 Workflow and Data Processing

To predict IAVs host tropism, we employed a 4-step workflow, as depicted in Figure 1. Firstly, we collected the genome data of IAVs and only retained high-quality sequences. Subsequently, we separated the genome data into training, validation, and test sets. Moreover, genome data were also encoded into amino acid sequences so that they can be processed by computers. Then, we used the constructed neural network of Flu-CNN for model training. Finally, we employed Flu-CNN to perform evaluations and further downstream analysis. This workflow enabled us to predict IAVs host tropism rapidly and accurately.

**Fig. 1.**
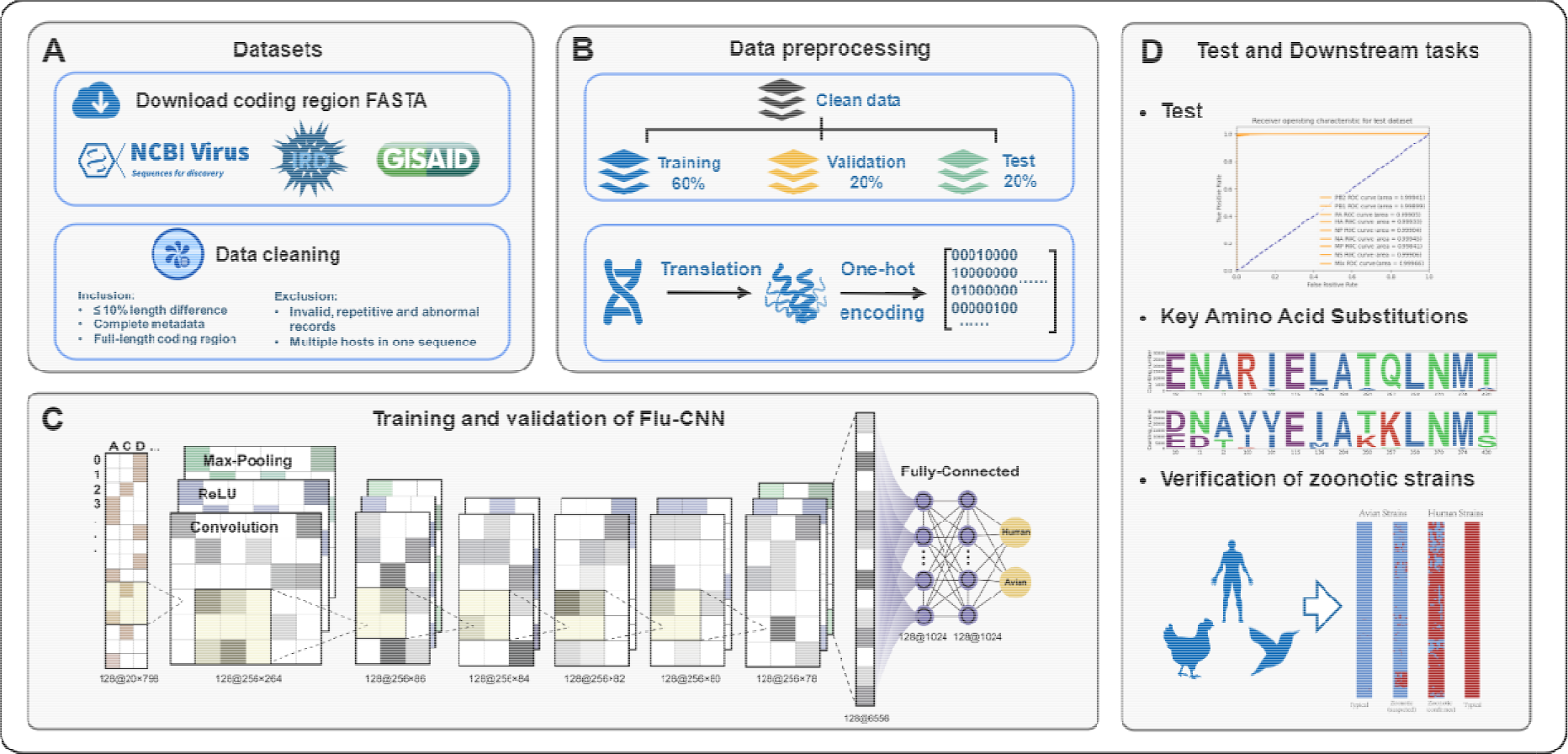
The workflow of IAVs host tropism prediction. The workflow is designed from left to right as follows: A. Data downloading and cleaning to generate datasets. B. Data preprocessing to partition datasets, translation and coding. C. Flu-CNN construction and training. D. Tropism prediction and downstream tasks, including predicting IAVs host tropism, screening key amino acid substitutions, and predicting zoonotic strains.

To obtain high-quality IAV genomic sequences, we retrieved RNA FASTA sequences of IAVs whole genome coding region from three major databases, including NCBI Virus, IRD and GISAID, as of October 14, 2022. As these databases contain numerous duplicate entries, we discarded strains with ambiguous characters, mislabeled epidemiological information, and incomplete metadata, and only retained one strain with consistent strain name and sequence. The reference strain (accession number A/New York/392/2004) on the NCBI Virus was used as the baseline, and only sequences within 10% difference in length were retained, and other sequences considered outliers (too long or too short) were discarded. Meanwhile, only sequences with hosts of human and avians are retains, with sequences of other hosts discarded. Consequently, a total of 911,098 sequences of 156,671 strains were obtained, including 630,656 sequences of 78,832 strains with the whole genome of eight segments. The sample distribution of these strains by host, subtype, year, and geographic region is presented in the supplementary material. We divided the genome data into training, validation, and test sets by a ratio of 6:2:2.

To enable the neural network model to recognize protein sequences, we use a unique one-hot encoding method that transforms each protein sequence into a matrix of values, where represents the type of amino acid and represents the length of the protein sequence. Each amino acid is represented by a particular row in the matrix. For instance, a sequence of amino acids in length would become a rectangular matrix of after unique thermal encoding; for the -th column, the first position is if the -th residue in the sequence is Alanine, and the rest positions are all.

### 2.2 Flu-CNN Structure and Model Training

Besides the size of the training data, the parameter size is also important to models, which determines whether the model can fit complex real-world scenarios. Convolutional Neural Network (CNN) is a type of deep learning model which is commonly used in computer vision and natural language processing, such as image recognition and object detection [38]. CNN models can extract local features from inputs like images or texts. Among them, the Char-CNN model has a simple structure with high accuracy and efficiency, making it suitable for text classification tasks. The basic structure of Char-CNN consists of two kinds of layers: a convolutional layer and a fully connected layer. The output of the pooling layer summarizes the input data to some extent. The fully connected layer uses the features extracted by convolution and pooling to output classification results, which uses the Softmax function as the activation function to normalize the predicted probability of each category.

Inspired by Char-CNN, we construct a deep network called Flu-CNN. It comprises six convolutional layers and three fully connected layers, with ReLU and Pooling in the convolutional layers and ReLU and Dropout in the fully connected layers [39, 40]. The final output is a two-dimensional vector, indicating the possibility of viral human/avians tropism. ReLU introduces nonlinearity, which addresses the gradient disappearance problem and reduces the dependency between neurons. Dropout is a regularization method that randomly discards some of the neurons in the neural network. Dropout can prevent the network from becoming too dependent on specific local features and can learn more robust features, which improves the performance on new samples.

In this study, we set the training epoch to 200, with a batch size of 128. The cross-entropy is used as the loss function, as shown in the following equation:

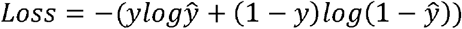

where *y* represents the true label, which takes the value of 0 or 1, and *ŷ* is the predicted label, which indicates the probability that the sample belongs to the positive case and takes the value from 0 to 1. The above equation is equivalent to -*logŷ* when *y* = 1 and -log(1 -*ŷ*), when *y* = 0. For a binary classification problem, the loss function converges to 0 if the model predicts correctly (*ŷ* is close to the true label value), and increases otherwise.

To evaluate the model, four metrics are taken into account, including accuracy, precision, recall, and F1-score, which are calculated as follows:

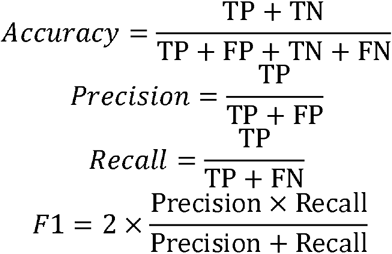

The above metrics are computed based on True Positive (TP), True Negative (TN), False Positive (FP), and False Negative (FN). TP represents the number of samples for which the classifier predicts positive cases as positive cases; TN is the number of samples for which the classifier predicts negative cases as negative cases; FP denotes the number of samples for which the classifier predicts negative cases as positive cases; and FN represents the number of samples for which the classifier predicts positive cases as negative cases.

The model was trained based on a specific given segment, or the whole genome as a conjunction of all segments. We selected the model parameter with the highest accuracy in the validation set throughout the training cycle as the final weights for each segment model. Once the training was completed, we used the best performing model weights to predict the test set.

### 2.3 Included Methods for Comparisons

We compared Flu-CNN with several state-of-the-art studies for a comprehensive study on our performance on predicting influenza viruses host tropism.

VIrus Deep learning HOst Prediction (VIDHOP) is a fast and accurate deep learning approach used for viral host prediction [41]. It requires partial sequences of the viral genome (100–400 bp long) without other virus features and predicts hosts at the species level for three viruses (IAVs, rabies hemolytic virus, and rotavirus A. VIDHOP can predict up to 36 host types for IAVs, 32 of which are closely related avian species. The architecture of VIDHOP for IAVs consists of three bidirectional Long short-term memory (LSTM) layers and two fully connected layers.

ML-(d)nts is a machine learning model used for predicting the nucleotide compositions of human-adapted IAVs [19]. Nucleotide compositions includes characterized mononucleotides (nts) and dinucleotides (dnts). The human adaptation of IAVs sequences were predicted by computing (d)nts features of six viral gene segments. The principal components analysis (PCA) and hierarchical clustering analysis revealed the linear separability of optimized (d)nts between the human-adaptive and avian-adaptive sets. The confusion matrix results and the area under the receiver operating characteristic curve indicate that the machine learning model has high performance in predicting human tropism of IAVs.

FluPhenotype is an online platform for early warning of IAVs, which accepts complete or partial genomic sequences to determine the virus phenotype rapidly [25]. An extensive collection of identified influenza virus molecular markers is available in FluPhenotype. Analysis of these molecular markers enables integrated inference of potential hosts of the viruses, including host type (avian, human, swine and other mammals), detailed host species, and probabilities for each host type. This method can be used for rapid determination of IAVs hosts, antigenicity, virulence, and drug resistance.

In addition, hosts of viral strains can be determined based on the phylogenetic tree of IAVs, which serves as a supplementary validation. The evolutionary tree of IAVs reveals different strains and their evolutionary relationships. Different epitope structures carried by different strains cause differences in host affinity, transmission ability, and so on. Information about the transmission paths, evolutionary patterns, and related characteristics of IAVs in different regions and time periods can also be revealed in the evolutionary tree.

## 3 Results

### 3.1 Host Tropism Prediction

#### 3.1.1 General Prediction of Host Tropism

To investigate the effectiveness of our approach, we compared Flu-CNN with two other methods, i.e., ML-(d)nts and VIDHOP, on a test set comprising 16,001 viral genomes that contained various subtypes. FluPhenotype and phylogenetic methods are not included because they cannot support a large number of strains. Phylogenetic presents challenges in constructing trees and distinguish virus host at a significant scale, and FluPhenotype requires individual genome-level operations on an online site. Table 1 presents the performance of studied methods, in which Flu-CNN outperformed other methods, both in individual gene segments and in the whole genome. In particular, our model achieved scores over 99% across all metrics for PB2, PA, HA, and the whole genome. On average, Flu-CNN outperforms VIDHOP by 9% in Recall and Accuracy, and by 5% in F1-score. And all four performance parameters were better than ML-(d)nts.

**Table 1.** Performance of Flu-CNN and compared methods on the test set. Because the ML-(d)nts method is not recommended for MP segment and NS segment, the corresponding result is not applicable (NA).

| Segment | PB2 |  |  | PB1 |  |  | PA |  |  |
| --- | --- | --- | --- | --- | --- | --- | --- | --- | --- |
| Method | Flu-CNN | ML-(d)nts | VIDHOP | Flu-CNN | ML-(d)nts | VIDHOP | Flu-CNN | ML-(d)nts | VIDHOP |
| Accuracy | 0.9963 | 0.9828 | 0.8253 | 0.9838 | 0.9781 | 0.7735 | 0.9908 | 0.9585 | 0.9116 |
| **Precision** | 0.9963 | 0.9836 | 0.9863 | 0.9839 | 0.9794 | 0.9859 | 0.9907 | 0.9628 | 0.9864 |
| **Recall** | 0.9963 | 0.9828 | 0.8253 | 0.9838 | 0.9781 | 0.7735 | 0.9908 | 0.9585 | 0.9116 |
| **F1-score** | 0.9963 | 0.9829 | 0.8950 | 0.9837 | 0.9783 | 0.8586 | 0.9907 | 0.9592 | 0.9465 |
| **Segment** | **HA** |  |  | **NP** |  |  | **NA** |  |  |
| **Method** | **Flu-CNN** | **ML-(d)nts** | **VIDHOP** | **Flu-CNN** | **ML-(d)nts** | **VIDHOP** | **Flu-CNN** | **ML-(d)nts** | **VIDHOP** |
| **Accuracy** | 0.9928 | 0.9819 | 0.9801 | 0.9850 | 0.9843 | 0.9350 | 0.9898 | 0.9816 | 0.9783 |
| **Precision** | 0.9928 | 0.9829 | 0.9867 | 0.9850 | 0.9849 | 0.9864 | 0.9898 | 0.9825 | 0.9868 |
| **Recall** | 0.9928 | 0.9819 | 0.9801 | 0.9850 | 0.9843 | 0.9350 | 0.9898 | 0.9816 | 0.9783 |
| **F1-score** | 0.9928 | 0.9821 | 0.9832 | 0.9849 | 0.9844 | 0.9592 | 0.9897 | 0.9818 | 0.9823 |
| **Segment** | **MP** |  |  | **NS** |  |  | **All Segments** |  |  |
| **Method** | **Flu-CNN** | **ML-(d)nts** | **VIDHOP** | **Flu-CNN** | **ML-(d)nts** | **VIDHOP** | **Flu-CNN** | **ML-(d)nts** | **VIDHOP** |
| **Accuracy** | 0.9861 | NA | 0.7600 | 0.9886 | NA | 0.9543 | 0.9955 | 0.9819 | 0.9775 |
| **Precision** | 0.9865 | NA | 0.9860 | 0.9886 | NA | 0.9866 | 0.9955 | 0.9830 | 0.9869 |
| **Recall** | 0.9861 | NA | 0.7600 | 0.9886 | NA | 0.9543 | 0.9955 | 0.9819 | 0.9775 |
| **F1-score** | 0.9862 | NA | 0.8486 | 0.9885 | NA | 0.9697 | 0.9955 | 0.9821 | 0.9819 |

In summary, our results demonstrate that Flu-CNN exhibits superior performance compared to state-of-the-art methods in predicting the host tropism of IAVs.

#### 3.2.1 Performance across Different Subtypes

Further, we explore different performance of each method on different subtypes. The test set is separated according to the subtype and the performance is further evaluated on this dimension. The performance on different subtypes is shown in Figure 2. Those approaches can all perform well on some subtypes, such as H2N1 and H3N2. However, in term of subtypes such as H1H1, H2N2 and H7N9, those methods can have different performance: Flu-CNN still maintains high accuracy, but the accuracy of other methods is limited. This shows that Flu-CNN not only has the best overall accuracy, but also achieves a high accuracy on individual subtypes. Hence, Flu-CNN exhibits its stability and performs the best across all subtypes. Such a stable performance across different subtypes of IAVs demonstrate the generality of our method in the field of IAV.

**Fig. 2.**
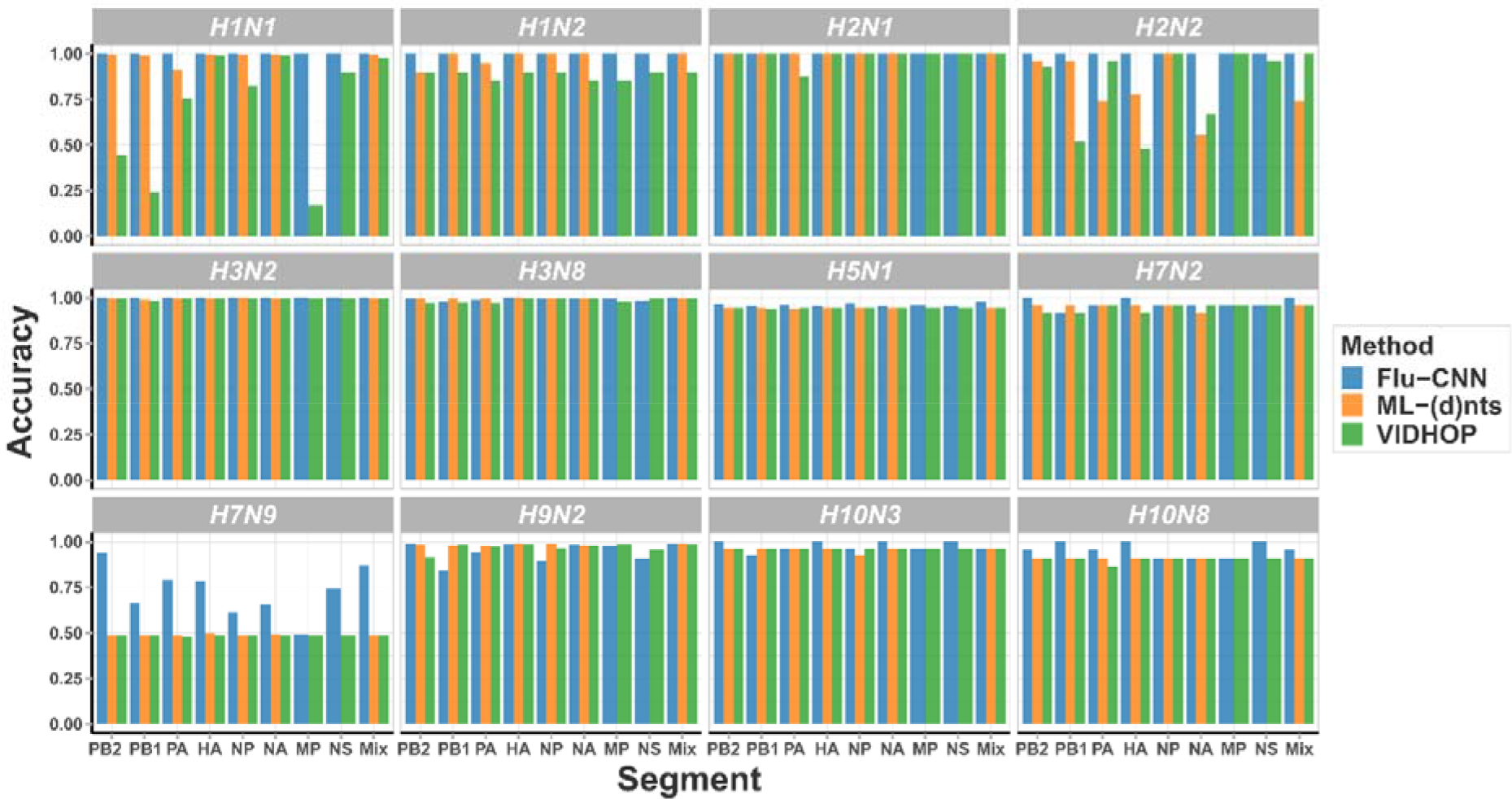
Histogram of accuracies on different subtypes. Each subtype is a subplot with horizontal coordinates indicating individual genome segments and genome-wide synthesis (Mix), and vertical coordinates indicating accuracy. Because the ML-(d)nts method is not recommended for MP segment and NS segment, the corresponding result is not applicable (NA).

#### 3.1.3 Specific Investigation on H5N1, H7N9, and H9N2

H3N2 and H1N1 are the predominating IAV subtypes, and they also account for the vast majority of our dataset. However, for some of the less abundant and minority subtypes, such as H5N1, H7N9, and H9N2, their insufficient data and significant bias may cause limited performance on these subtypes. Despite minority, they are the top three most infected human in avian influenza. So, this subsection particularly investigates the performance on those subtypes. And all the four state-of-the-art methods, ML-(d)nts and, VIDHOP, we have added other two methods, phylogenetic and FluPhenotype, and the phylogenetic method, are included, because the experiment is conducted on small-scale dataset.

All sequences of the above three subtypes, including the training set, validation set and test set, are analyzed and compared using Flu-CNN and other methods. We randomly sampled 100 sequences from three subtypes by the ratio of human to avian 1:1 for 20 times, as the dataset for performance evaluation. In this subsection, the experiment is conducted on small-scale dataset, so all the four state-of-the-art methods, ML-(d)nts, VIDHOP, FluPhenotype, and the phylogenetic method, are included.

The scatter plots of sampling accuracy of different methods on H5N1, H7N9, and H9N2 subtypes are shown in Figure 3. It can be observed that VIDHOP and ML-(d)nts methods may have difficulty in identifying the host species in these three subtypes, with the accuracy only around 50%. Although the phylogenetic and FluPhenotype may be unstable in accuracy, they performed better compared to the VIDHOP and ML-(d)nts methods. Among all the five methods, our Flu-CNN still achieves the best performance, presenting both the most stable and highest accuracy across all three subtypes.

**Fig. 3.**
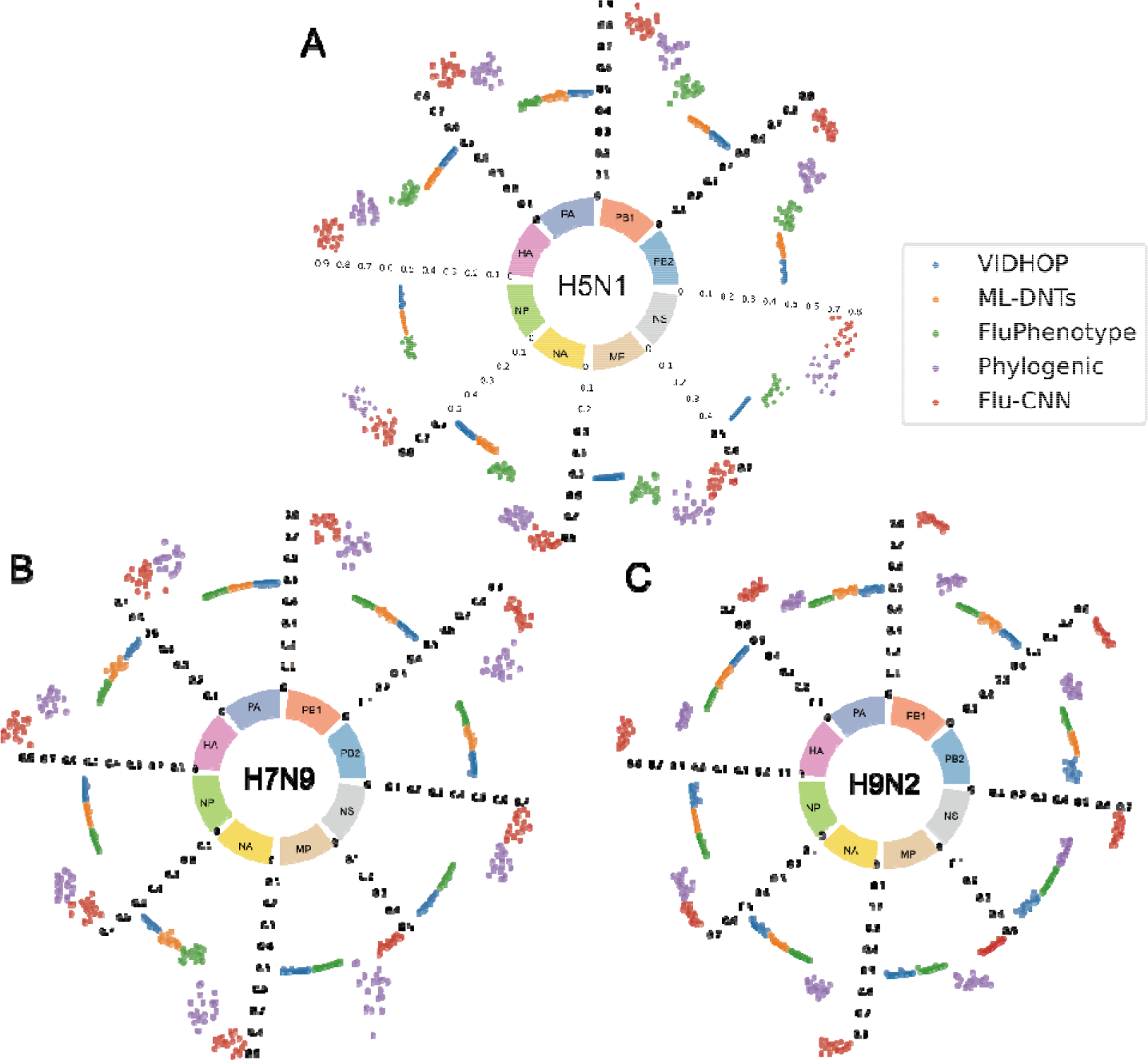
Ring bar chart of accuracy on certain subtypes: A. H5N1. B. H7N9. C. H9N2. Each sector area represents a genomic segment. Each point represents one sample of accuracy result of methods: the blue dots for VIDHOP, the orange dots for ML-(d)nts method, the green dots for FluPhenotype method, the purple dots for phylogenic method, and the red dots for Flu-CNN.

### 3.2 Interpretability of Flu-CNN

Neural networks are often considered as a black box, and it is challenging to understand their underlying working mechanisms and internal computation process intuitively. In this subsection, we visualize the middle layer of Flu-CNN to understand the feature representation inside the model and investigate whether it learns the valuable feature information. We utilized Uniform Manifold Approximation and Projection (UMAP) to conduct the dimension reduction [24], by which we projected the middle layer vectors into a two-dimensional space for further visualizations.

To take HA and NA segments as examples, their UMAP visualizations are shown in Figure 4. Obviously, different host tropism can be clearly distinguished in the UMAP visualization, and therefore Flu-CNN can learn the key features to distinguish the hosts by convolution layers. In addition to hosts, Flu-CNN is also capable of extracting important features to distinguish subtypes. The UMAP visualization shows that sequences of different subtypes can be separated by the model. Consequently, the convolutional network of Flu-CNN not only extracts the host tropism information of IAVs, but also can support the classification of different subtypes.

**Fig. 4.**
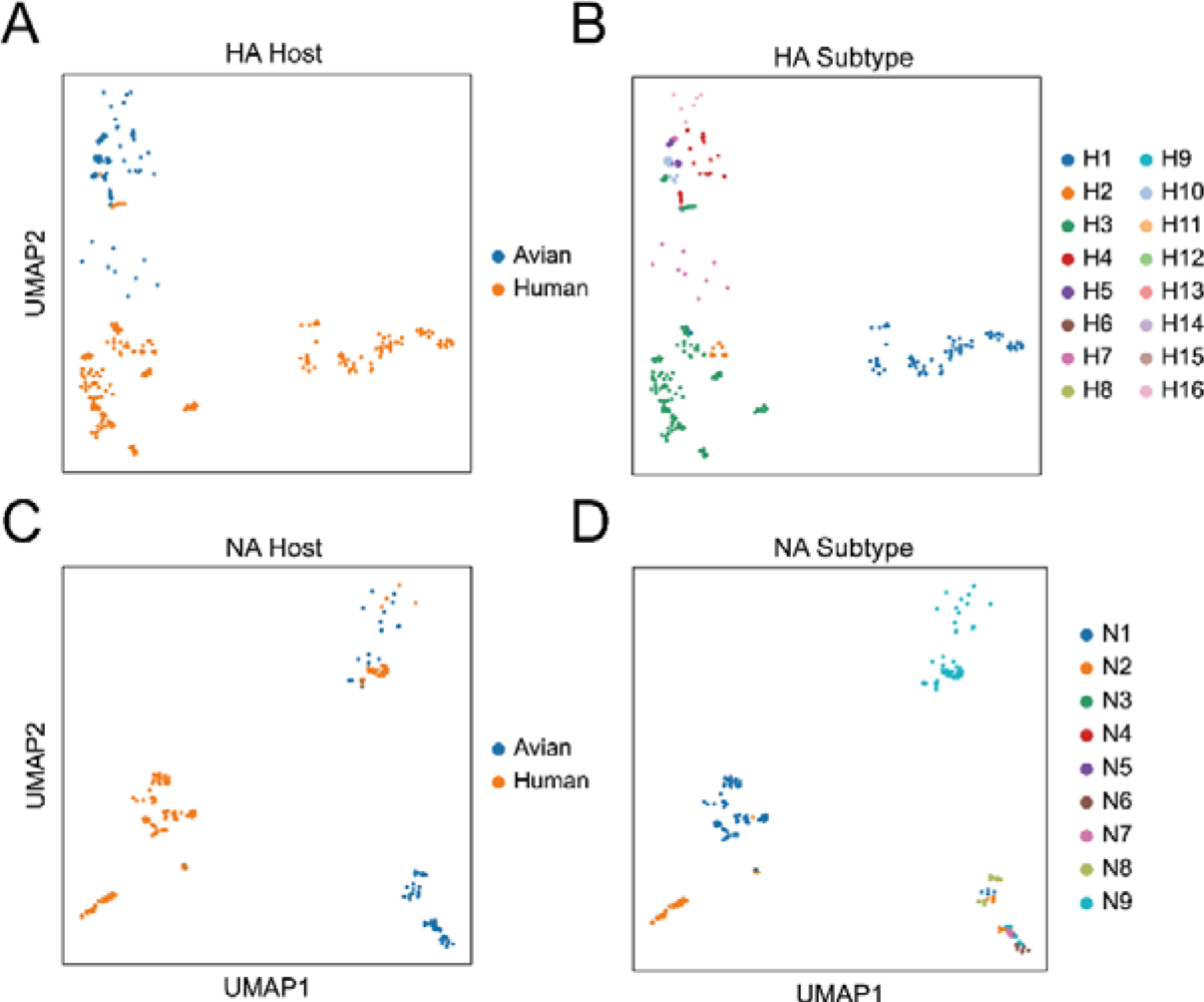
UMAP visualization of the convolutional layer output in Flu-CNN. Different hosts and subtypes are represented by different color points. A. HA segment colored by hosts. B. HA segment colored by subtypes. C. NA segment colored by hosts. D. NA segment colored by subtypes.

Notably, Flu-CNN has only six convolutional layers and three fully connected layers, with less than 10 million parameters. Compared with other large models with hundreds of layers and billions of parameters, our model may appear simple. Even so, Flu-CNN can still extract important features, and achieves remarkable performance in predicting the host tropism of IAV, which demonstrate the effectiveness of our method.

### 3.3 Identifying Key Amino Acid Substitutions for Host Tropism Transition of IAVs

The antigenicity of influenza virus proteins is an important factor in the host tropism [18]. Previous studies have detected numerous amino acid phenotypes as biomarkers of human-adapted IAVs, which plays a critical role in cross-host transmission of avian influenza [25]. Hence, identifying human adaptive amino acid phenotypes of influenza viruses is of great significance to the surveillance and pre-warning of the influenza. Conventionally, these phenotypes have been determined primarily through biological experiments, which can be generally accurate. However, experimental methods can be both time-consuming and labor-intensive, and further requires a laboratory of biosafety level 3 [26], which may be vastly expensive for large-scale studies.

From a computational perspective, such substitutions can be identified by Flu-CNN. Specifically, we can examine individual substitutions respectively, by using Flu-CNN to investigate the change of host tropism after applying mutations to the gene segment. Thus, Flu-CNN can effectively identify specific amino acid mutations by estimating the effect of each mutation on the IAV host tropism. With a focus on PB2, PA, and NP, we have identified several key amino acid substitutions affecting human tropism of avian influenza viruses.

To take the PB2 protein as an example, Flu-CNN screened eight important amino acid substitutions (T108V, A274S, S286G, Q591R, Q591K, E627K, D701N, D701E) for host adaptability. Figure 5 visualizes these eight substitutions in the visualized structure of PB2 protein. It can be found that these substitutions are all located on the outer surface of the protein. Of these, five mutations (S286G, Q591R, Q591K, E627K, D701N) have been biologically validated as key amino acid phenotypes for human tropism of IAV, by current literatures [27-29]. The other three substitutions (T108V, A274S, D701E) are also located in important functional regions. The T108V mutation is at the N-terminal of PB2 protein, in the minimal recognition sequence for the binding of PB1 protein and NP protein in the polymerase heterotrimer. A274S is at the N-terminal of PB2 protein, in the sequence associated with cap binding. D701E is at the C-terminal of PB2 protein, in the same position as the D701N substitution. Considering their structural functions, it can be concluded that they may play a part in the host tropism, although their detailed effects still remain to be elucidated in future investigations. Results and visualizations for PA and NP segments are presented in Supplementary Material.

**Fig. 5.**
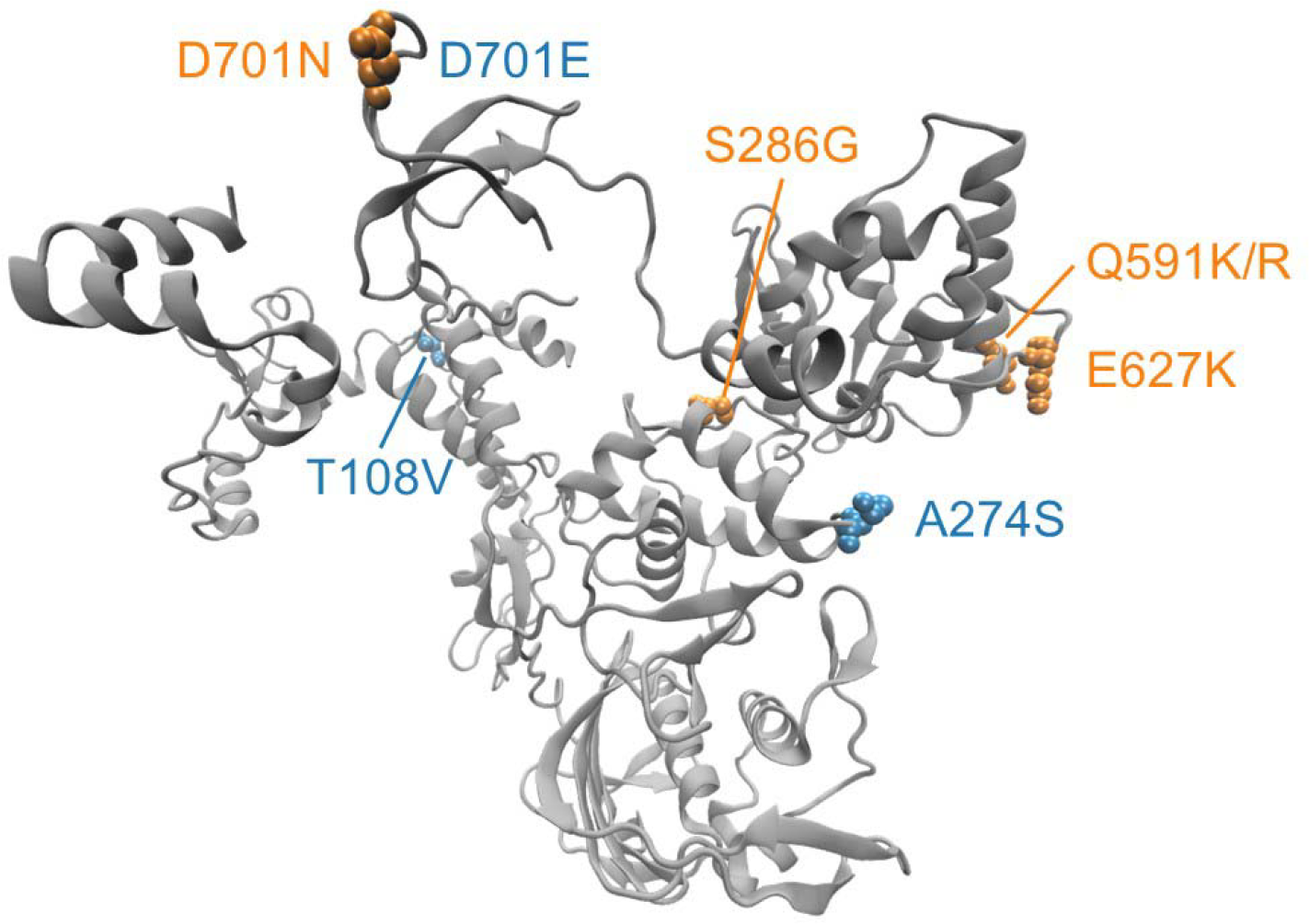
The key human-adapted amino acid substitutions of PB2 protein (PDB: 6QPF) [34] screened by Flu-CNN, visualized by Visual Molecular Dynamics (VMD) [35, 36]. The selected amino acid substitutions are denoted in blue and yellow. Yellow indicates that the substitution has been experimentally verified, and blue indicates that the substitution has not been reported in the current literature, with other areas in grey.

### 3.4 Verification on Zoonotic IAV Strains

Zoonotic IAVs can pose a significant threat, which may bring about global epidemics. In case of such a threat, Flu-CNN can serve for the identification and prediction of zoonotic strains. Christine L. P. Eng have retrieved and studied discriminating zoonotic strains in four groups, including 5,685 typical avian influenza strains, 5,110 human typical influenza strains, 126 confirmed zoonotic influenza strains of human origin, and 346 suspected zoonotic influenza strains of avian origin [16]. Based on those strains, we employed our model to categorize their host tropism, which is colored by hosts and visualized in groups.

The categorized zoonotic IAVs are visualized in Figure 6. As shown in Figure 6, most of the IAVs had only a single host tropism. Both the typical avian strains in Figure 6A and human strains in Figure 6D demonstrate the uniformity in host tropism. In contrast, the suspected zoonotic strains isolated from avian sources (Figure 6B) and confirmed zoonotic strains isolated from human sources during zoonotic outbreaks (Figure 6C) displayed a mosaic mixing pattern in their genomic segments. Among the confirmed zoonotic strains, the proportion of human tropic strains was significantly higher than that of suspected zoonotic strains. This phenomenon shows that these strains do have some zoonotic risk and the risk of confirmed zoonotic strains is higher than that of suspected zoonotic strains.

**Fig. 6.**
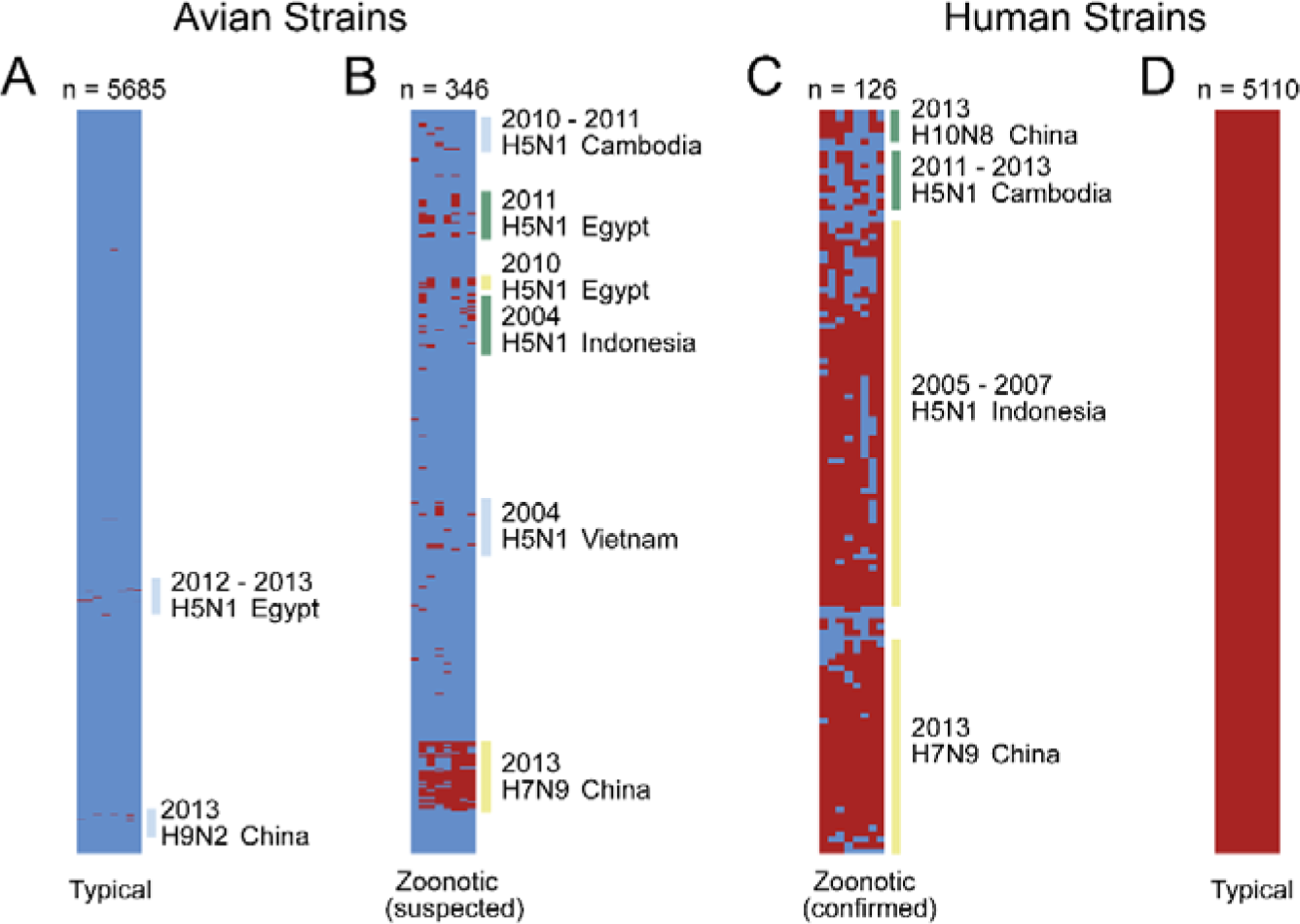
Segmental host tropism signatures of human, avian and zoonotic strains from Christine. Each row represents a strain, and each column represents a gene segment, with red representing human adaption and blue representing avian adaption. A. Typical avian strain. B. Suspected zoonotic strains isolated from avian during zoonotic outbreaks. C. Confirmed zoonotic strains isolated from human during zoonotic outbreaks. D. Typical human strain.

Notably, the result of Flu-CNN is consistent with the work of Christine. This indicates that the species barrier does exist between various classes of influenza virus host, which prevents most avian Influenza viruses with only avian genes from free cross-host transmissions.

Furthermore, we utilized Flu-CNN to identify zoonotic strains in the entire dataset collected in this research. As shown in Figure 7, the vast majority of strains are single host-adapted, which is incapable of cross-host transmissions. However, there are seven lineages that may have zoonotic risks, which show a mosaic pattern of host adaptability. These strains cover four subtypes of H5N1, H5N6, H7N9 and H9N2, as depicted in Figure 7.

**Fig. 7.**
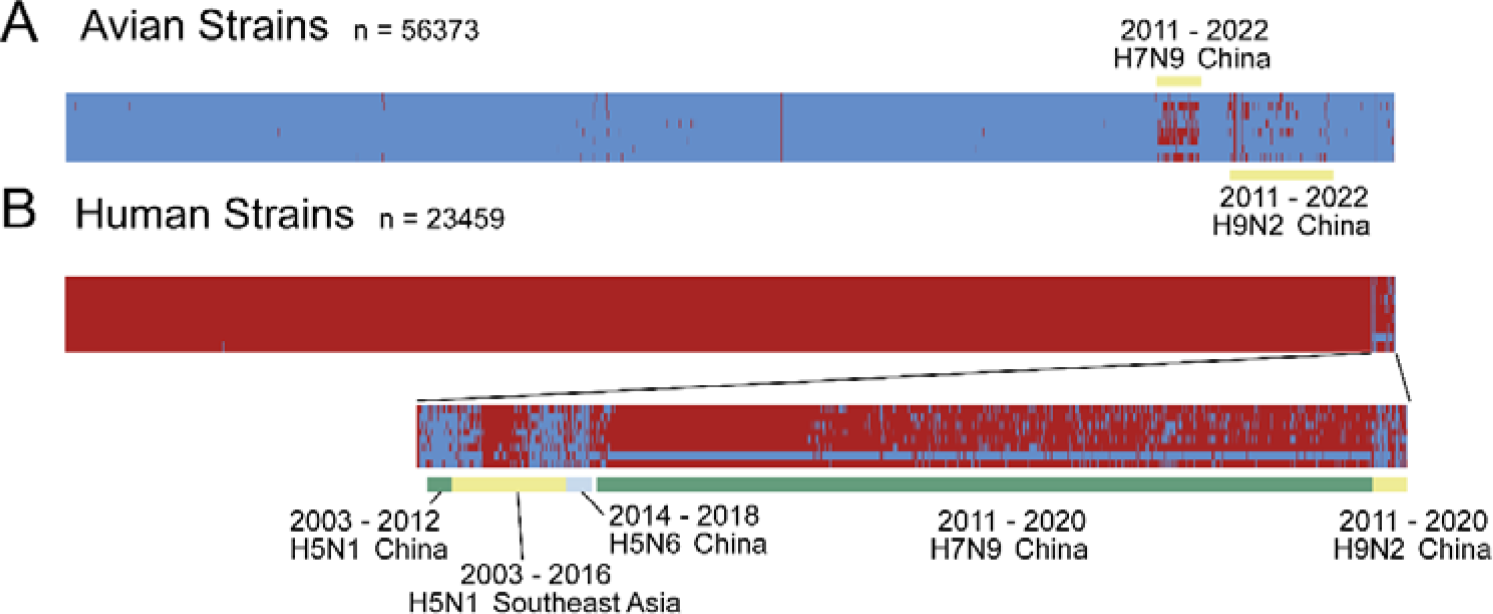
Segmental host tropism signatures of human, avian and zoonotic strains from the entire data in this research. Each column represents a strain, and each row represents a gene segment, with red representing human adaption and blue representing avian adaption. A. Avian strains. B. Human strains.

## 4 Discussion

The host that IAV can infect is strictly limited by its host tropism, and changes in host tropism may lead to cross-host transmission. While numerous factors can function in the viral host tropism, there is no systematic criterion for assessing those factors. Therefore, the identification of IAVs host tropism has been an important research issue. Meanwhile, deep learning has been widely applied in the fields of protein structure prediction, protein function prediction, and genetic engineering, and is vastly promising for the host tropism investigation of IAVs [30-32]. Benefitted from this, we used a powerful deep network to distinguish viral tropism in different hosts more effectively and efficiently.

This research focuses on analyzing patterns of IAV tropism in humans and avian species and establishing a method of rapid identification of IAV host tropism. We collected the largest dataset of IAV sequences to date, which is approximate to one million sequences. These sequences showed a clear bias, mostly for the H3N2 and H1N1 subtypes, originating from the United States and China. We constructed Flu-CNN, which demonstrated outstanding performance with an accuracy of over 99% in all segments of the genome. Compared to other methods, Flu-CNN exhibited exceptional stability and superior performance, especially for H5N1, H7N9, and H9N2 subtypes with cross-host transmission. Compared with Flu-CNN, other methods may have slightly weaker performance due to various reasons. In general, it could be that the IAVs data used has a significant bias. Specifically, the ML-(d)nts method innovatively proposes that nucleotide composition features are related to IAVs host adaptation, the VIDHOP method is better at identifying hosts at the species level, and the FluPhenotype method focuses on comprehensive analysis of IAVs phenotypes.

The better performance of Flu-CNN can be explained by various reasons, including the large size of training data, the sufficient number of training sessions, and the effective learning structure by our proposed model. Besides, our analysis of intermediate layer vectors generated by Flu-CNN indicated that the convolutional network could effectively extract high-dimensional information from genome sequences. It is generally believed that convolutional networks are effective in extracting local features of the input data. With more convolutional layers and pooling layers, convolutional networks can also contribute to the extraction of global features. Notably, the capability of our approach for distinguishing subtypes and establishing key viral features suggests that convolutional networks can reveal the underlying information within viral sequences and even accomplish additional tasks.

All methods show fluctuating performance on subtypes. The accuracy of different subtypes varies greatly, and the accuracy of some subtypes is relatively poor. There may be many reasons for this situation. Some subtypes have a small number of sequences, low richness within the sequence, and high sequence similarity between different subtypes. So far, how these subtypes spread across hosts and how to evolve in the next step have yet to be studied which demonstrate that we still know little about them. Accurate identification of different subtypes of host tropism is not a simple matter. Even so, our method can still effectively mine useful information and maintain high accuracy, which shows the effectiveness of our method.

Although there has been no clear evidence of direct human-to-human transmission of avian influenza viruses, the possibility for their evolving into human-to-human transmissible viruses cannot be utterly denied. Once the mutations or reassortment occurs in such viruses, it is likely that they can acquire the ability to transmit between human beings and therefore change the host tropism. Accordingly, it is crucial to identify human-adapted amino acid phenotypes in IAVs, which can significantly contribute to the study on antigenic epitope signatures and the assessment of viral risk. Nevertheless, such an identification solely based on experimental screening can be inefficient and resource-intensive. In the present study, we screened key amino acid substitutions of certain segmental proteins using Flu-CNN and found several important mutations. Most of the discovered substitutions can be validated to be effective by supportive literature references, which demonstrate the effectiveness of our screening. For those with no supportive references, they may serve as candidates for human-adapted amino acid substitutions, which serves as the guidance for future biological investigations.

While identifying human-adapted amino acid phenotypes, we take the PB2 protein as a major example, because it is indispensable to virus replication and is a pivotal determinant of host range [33]. Researchers have discovered that distinct PB2 proteins affect viral growth performance, pathogenicity, and infection range [34-36]. Moreover, PB2 proteins are implicated in signaling pathways that follow viral infection, including blocking JAK1/STAT signaling via targeting JAK1 for degradation through proteasomal mechanisms, indicating that the PB2 protein is essential in regulating the interaction between virus and host [37]. Meanwhile, we have also screened PA and NP, whose results are presented in Supplementary Material.

The avian influenza viruses that threaten humans are usually zoonotic influenza viruses. Human infections by those viruses are usually through direct contact with infected animals or contaminated environments, which do not spread from human to human. However, if these viruses acquire the capability of sustainable human-to-human transmission, they could cause a pandemic because humans have very limited immunity to them. Hence, the early detection and surveillance of zoonotic influenza viruses is vastly important. This study shows that Flu-CNN is capable of detecting avian influenza strains that may cross over from avian to human by identifying zoonotic strains. From the zoonotic results by Flu-CNN, the host tropism of each segment gene is mostly the same. Subtypes H5N1, H7N9, and H9N2 account for the majority of zoonotic strains, and geographically, China and Southeast Asia are frequent outbreaks, and these subtypes and regions should be the focus of our outbreak surveillance.

## 5 Conclusion

This paper has proposed a deep neural network approach named Flu-CNN as a valuable tool for identifying the host tropism of IAVs. Our approach can rapidly identify the host tropism of viruses merely by viral genomic sequences, without extracting any abstract features. It achieves more than 99 % accuracy and maintains its stability in accuracy, which enjoys the best performance across different gene segments and subtypes. The interpretability study demonstrate that our model can capture valuable features from genome sequences, and can even support the classification of subtypes. We have also used Flu-CNN to identify amino acid substitutions that affect host adaptability of IAV and to assess the zoonotic risk of viral strains.

In summary, this is a valuable approach for analyzing the potential risk and the genomic data of IAVs. This research also produces innovative insights into the evolutionary characterization of IAV, which may contribute to the surveillance of potential outbreaks and spread of IAVs.

## Data Availability

All data produced in the present study are available upon reasonable request to the authors

https://www.ncbi.nlm.nih.gov/labs/virus

## Highlights

- The proposed Flu-CNN is currently the most accurate method to predict IAV host tropism.
- Key amino acid substitutions that affect IAV host adaption can be identified by Flu-CNN.
- Flu-CNN can effectively predict the zoonotic risk of IAV strains.

## Acknowledgement

We gratefully acknowledge the scientific community on the NCBI VIRUS, IRD and GISAID platform and all contributing experts in influenza A virus sequences.

## Availability and lmplementation

All the data, source code and documentation are available at https://github.com/southwood-luo/Flu-CNN.

## References

1. Ren H, Jin Y, Hu M et al. Ecological dynamics of influenza A viruses: cross-species transmission and global migration, Sci Rep 2016;6:36839.

2. Gong X, Hu M, Chen W et al. Reassortment Network of Influenza A Virus, Frontiers in Microbiology 2021;12:793500.

3. Nuwarda RF, Alharbi AA, Kayser V. An Overview of Influenza Viruses and Vaccines, Vaccines (Basel) 2021;9.

4. Wille M, Barr IG. Resurgence of avian influenza virus, SCIENCE 2022;376:459–460.

5. Scarafoni D, Telfer BA, Ricke DO et al. Predicting Influenza A Tropism with End-to-End Learning of Deep Networks, Health Security 2019;17:468–476.

6. Long JS, Mistry B, Haslam SM et al. Host and viral determinants of influenza A virus species specificity, NATURE REVIEWS MICROBIOLOGY 2019;17:67–81.

7. Vijaykrishna D, Mukerji R, Smith GJ. RNA Virus Reassortment: An Evolutionary Mechanism for Host Jumps and Immune Evasion, PLoS Pathogens 2015;11:e1004902.

8. Nelson MI, Holmes EC. The evolution of epidemic influenza, NATURE REVIEWS GENETICS 2007;8:196–205.

9. Li KS, Guan Y, Wang J et al. Genesis of a highly pathogenic and potentially pandemic H5N1 influenza virus in eastern Asia, NATURE 2004;430:209–213.

10. Peiris M, Yuen KY, Leung CW et al. Human infection with influenza H9N2, LANCET 1999;354:916–917.

11. Li YT, Linster M, Mendenhall IH et al. Avian influenza viruses in humans: lessons from past outbreaks, Br Med Bull 2019;132:81–95.

12. WHO WHO (2023), ’Avian Influenza Weekly Update Number 884’.

13. Webster RG, Bean WJ, Gorman OT et al. Evolution and ecology of influenza A viruses, Microbiol Rev 1992;56:152–179.

14. Kislinger T, Cox B, Kannan A et al. Global survey of organ and organelle protein expression in mouse: combined proteomic and transcriptomic profiling, CELL 2006;125:173–186.

15. Bouvier NM. Animal models for influenza virus transmission studies: a historical perspective, Current Opinion in Virology 2015;13:101–108.

16. Eng CL, Tong JC, Tan TW. Distinct Host Tropism Protein Signatures to Identify Possible Zoonotic Influenza A Viruses, PLoS One 2016;11:e150173.

17. Eng C, Tong JC, Tan TW. Predicting Zoonotic Risk of Influenza A Viruses from Host Tropism Protein Signature Using Random Forest, INTERNATIONAL JOURNAL OF MOLECULAR SCIENCES 2017;18.

18. Qiang X, Kou Z, Fang G et al. Scoring Amino Acid Mutations to Predict Avian-to-Human Transmission of Avian Influenza Viruses, MOLECULES 2018;23.

19. Li J, Zhang S, Li B et al. Machine Learning Methods for Predicting Human-Adaptive Influenza A Viruses Based on Viral Nucleotide Compositions, MOLECULAR BIOLOGY AND EVOLUTION 2020;37:1224–1236.

20. Zhang X, Zhao J, LeCun Y (2015), ’Character-level convolutional networks for text classification’, Proceedings of the 28th International Conference on Neural Information Processing Systems - Volume 1, MIT Press, Montreal, Canada, pp. 649-657.

21. Hatcher EL, Zhdanov SA, Bao Y et al. Virus Variation Resource - improved response to emergent viral outbreaks, NUCLEIC ACIDS RESEARCH 2017;45:D482–D490.

22. Shu Y, McCauley J. GISAID: Global initiative on sharing all influenza data - from vision to reality, Euro Surveill 2017;22.

23. Zhang Y, Aevermann BD, Anderson TK et al. Influenza Research Database: An integrated bioinformatics resource for influenza virus research, NUCLEIC ACIDS RESEARCH 2017;45:D466–D474.

24. McInnes L, Healy J. UMAP: Uniform Manifold Approximation and Projection for Dimension Reduction, ArXiv 2018;abs/1802.03426.

25. Lu C, Cai Z, Zou Y et al. FluPhenotype-a one-stop platform for early warnings of the influenza A virus, BIOINFORMATICS 2020;36:3251–3253.

26. Radigan KA, Misharin AV, Chi M et al. Modeling human influenza infection in the laboratory, Infection and Drug Resistance 2015;8:311–320.

27. Wen L, Chu H, Wong BH et al. Large-scale sequence analysis reveals novel human-adaptive markers in PB2 segment of seasonal influenza A viruses, Emerg Microbes Infect 2018;7:47.

28. Yamada S, Hatta M, Staker BL et al. Biological and structural characterization of a host-adapting amino acid in influenza virus, PLoS Pathogens 2010;6:e1001034.

29. Manz B, de Graaf M, Mogling R et al. Multiple Natural Substitutions in Avian Influenza A Virus PB2 Facilitate Efficient Replication in Human Cells, JOURNAL OF VIROLOGY 2016;90:5928–5938.

30. Kim H, Park K, Yon JM et al. Predicting multipotency of human adult stem cells derived from various donors through deep learning, Sci Rep 2022;12:21614.

31. Guzzi PH, Lomoio U, Puccio B et al. Structural analysis of SARS-CoV-2 Spike protein variants through graph embedding, Netw Model Anal Health Inform Bioinform 2023;12:3.

32. Wang H, Ma X. Learning discriminative and structural samples for rare cell types with deep generative model, BRIEFINGS IN BIOINFORMATICS 2022;23.

33. Subbarao EK, London W, Murphy BR. A single amino acid in the PB2 gene of influenza A virus is a determinant of host range, JOURNAL OF VIROLOGY 1993;67:1761–1764.

34. Kim G, Shin HM, Kim HR et al. Effects of host and pathogenicity on mutation rates in avian influenza A viruses, Virus Evol 2022;8:c13.

35. Ivan FX, Kwoh CK. Rule-based meta-analysis reveals the major role of PB2 in influencing influenza A virus virulence in mice, BMC GENOMICS 2019;20:973.

36. Choi EJ, Lee YJ, Lee JM et al. The effect of mutations derived from mouse-adapted H3N2 seasonal influenza A virus to pathogenicity and host adaptation, PLoS One 2020;15:e227516.

37. Yang H, Dong Y, Bian Y et al. The influenza virus PB2 protein evades antiviral innate immunity by inhibiting JAK1/STAT signalling, Nature Communications 2022;13:6288.

38. LeCun Y, Bengio Y, Hinton G. Deep learning, NATURE 2015;521:436–444.

39. Glorot X, Bordes A, Bengio Y. Deep Sparse Rectifier Neural Networks, JOURNAL OF MACHINE LEARNING RESEARCH 2011;15:315–323.

40. Srivastava N, Hinton G, Krizhevsky A et al. Dropout: A Simple Way to Prevent Neural Networks from Overfitting, JOURNAL OF MACHINE LEARNING RESEARCH 2014;15:1929–1958.

41. Mock F, Viehweger A, Barth E et al. VIDHOP, viral host prediction with deep learning, BIOINFORMATICS 2021;37:318–325.

