## Supplemental Material for "Flu-CNN: predicting host tropism of influenza A viruses via character-level convolutional networks"

### 1 Supplementary Figures

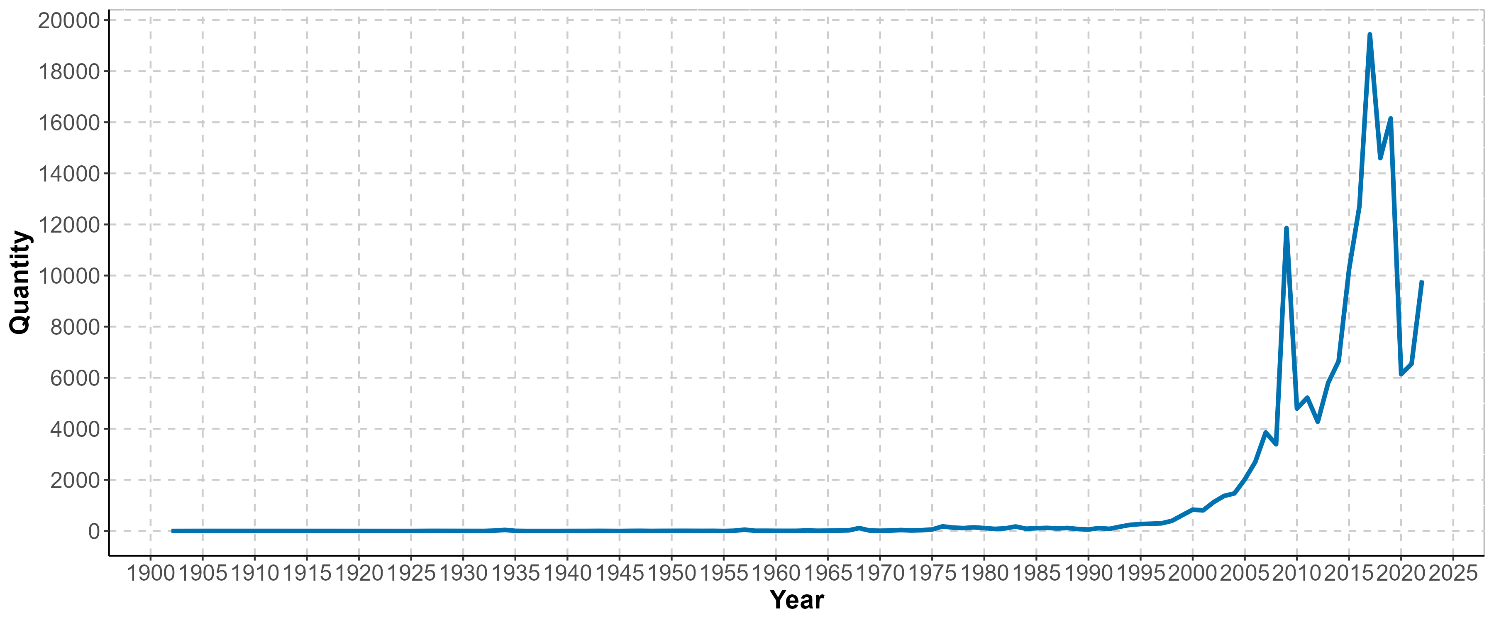

**Supplementary Figure 1.** Temporal distribution of metadata. The horizontal coordinate represents the year and the vertical coordinate represents the number.

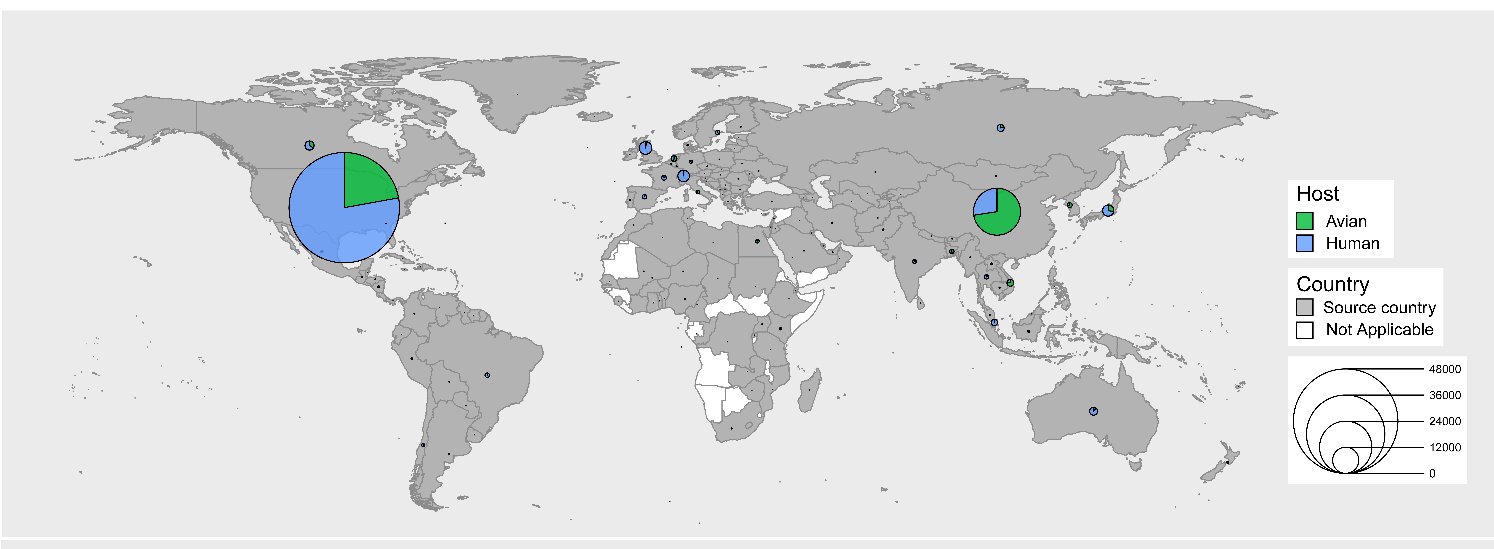

**Supplementary Figure 2.** Geographical distribution of metadata. The pie chart size represents the number, the gray area represents the source country, and the white area represents the sample that was not collected for that region.

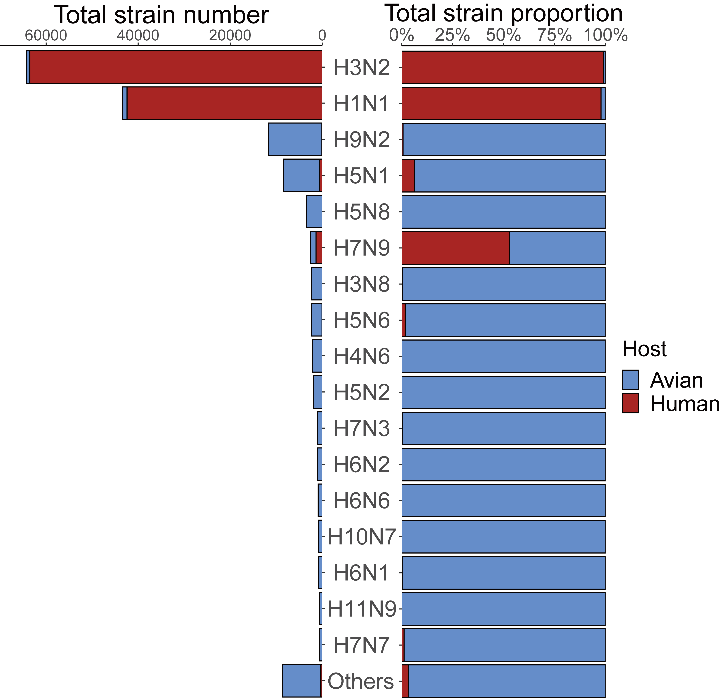

**Supplementary Figure 3.** Subtype distribution of metadata. The left side represents the number of different subtypes, and the right side represents the proportion after normalization of the number.

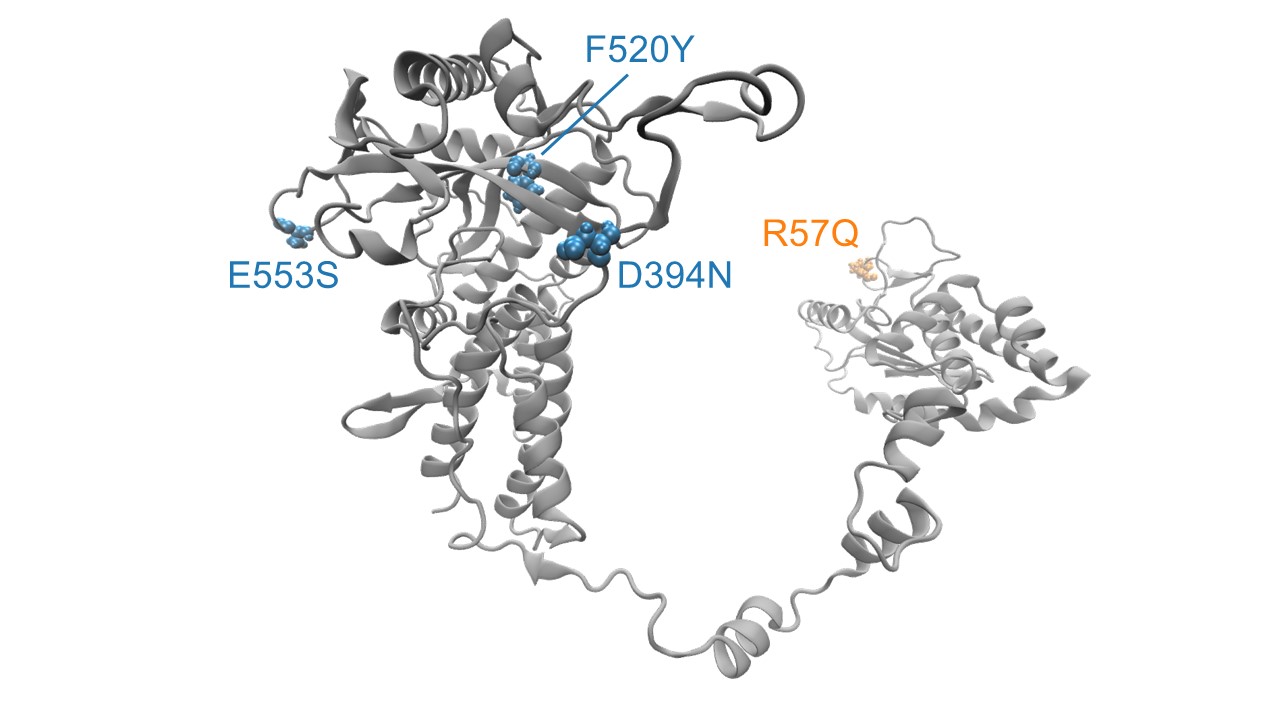

**Supplementary Figure 4.** The key human-adapted amino acid substitutions of PA protein (PDB: 7NHC) [1] screened by Flu-CNN, visualized by Visual Molecular Dynamics (VMD) [2, 3]. The selected amino acid substitutions are denoted in blue and yellow. Yellow indicates that the substitution has been experimentally verified [4, 5], and blue indicates that the substitution has not been reported in the current literature, with other areas in grey.

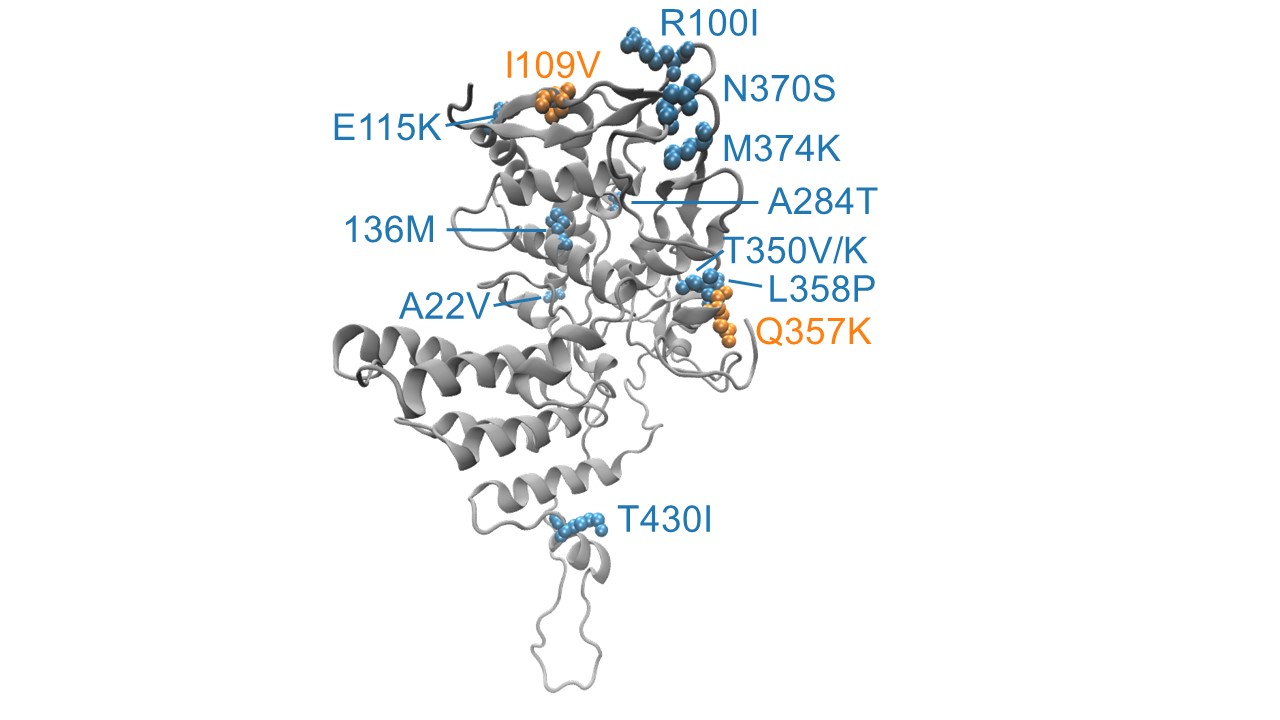

**Supplementary Figure 5.** The key human-adapted amino acid substitutions of NP protein (PDB: 7Q06) [6] screened by Flu-CNN, visualized by VMD. The selected amino acid substitutions are denoted in blue and yellow. Yellow indicates that the substitution has been experimentally verified [4, 5], and blue indicates that the substitution has not been reported in the current literature, with other areas in grey.

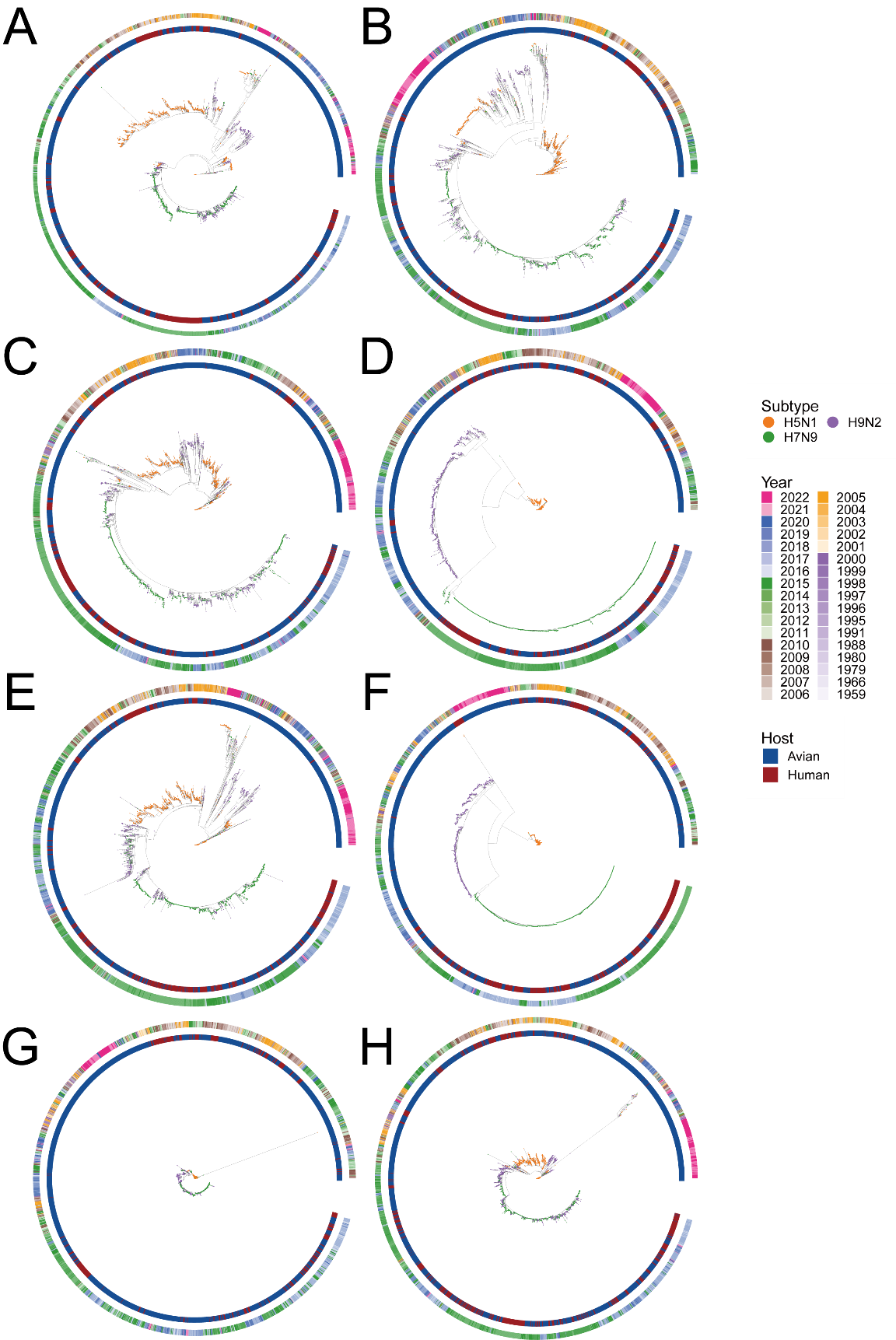

**Supplementary Figure 6.** The phylogenetic tree of H5N1, H7N9, and H9N2 visualized by ggtree [7]. The points on the phylogenetic tree represent the subtypes of the strain, the inner ring represents the host of the strain, and the outer ring represents the year of collection of the strain. Each subplot represents an IAV segment. A. PB2. B. PB1. C. PA. D. HA. E. NP. F. NA. G. MP. H. NS.

**References:**

[1].	Keown, J.R., et al., Mapping inhibitory sites on the RNA polymerase of the 1918 pandemic influenza virus using nanobodies. Nat Commun, 2022. 13(1): p. 251.

[2].	Humphrey, W., A. Dalke and K. Schulten, VMD: visual molecular dynamics. J Mol Graph, 1996. 14(1): p. 33-8, 27-8.

[3].	Stone, J.E., K.L. Vandivort and K. Schulten. GPU-accelerated molecular visualization on petascale supercomputing platforms. in Proceedings of the 8th International Workshop on Ultrascale Visualization. 2013. Denver, Colorado: Association for Computing Machinery.

[4].	Finkelstein, D.B., et al., Persistent host markers in pandemic and H5N1 influenza viruses. J Virol, 2007. 81(19): p. 10292-9.

[5].	Chen, G.W., et al., Genomic signatures of human versus avian influenza A viruses. Emerg Infect Dis, 2006. 12(9): p. 1353-60.

[6].	Kincannon, W.M., et al., Biochemical and structural characterization of an aromatic ring-hydroxylating dioxygenase for terephthalic acid catabolism. Proc Natl Acad Sci U S A, 2022. 119(13): p. e2121426119.

[7].	Guangchuang, et al., ggtree: an r package for visualization and annotation of phylogenetic trees with their covariates and other associated data. Methods in Ecology & Evolution, 2017.
